# Global research trends in postoperative ileus from 2011 to 2023: A Scientometric study

**DOI:** 10.1101/2024.01.12.24301260

**Authors:** Yan Zhou, Zi-Han Yin, Ming-Sheng Sun, Yang-Yang Wang, Chen Yang, Shu-Hao Li, Fan-Rong Liang, Fang Liu

## Abstract

**Objective:** To explore the current status, trends, and frontiers of POI research from 2011 to the present based on bibliometric analysis.

**Methods:** Publications published on POI research from 2011 to 2023 were retrieved on June 1, 2023, from the Web of Science Core Collection. CiteSpace 6.2.R2 and VOSviewer were used to conduct bibliometric visualization.

**Results:** In total, 778 POI records published from 2011 to 2023 were retrieved. Over the past few decades, the annual cumulative number of related articles has linearly increased, with China and the United States of America (USA) contributing prominently. All the publications were from 59 countries and territories. China and the University of Bonn were the top contributing country and institution, respectively. *Neurogastroenterology & Motility* was the most prolific journal. The Journal of *Gastrointestinal Surgery* had the highest number of citations. Wehner Sven was the most productive author. Burst keywords (*e.g.*, colon, prolonged ileus, acupuncture, paralytic ileus, pathophysiology, rectal cancer, gastrointestinal function, risk) and a series of reference citation bursts provided evidence for the research frontiers in recent years.

**Conclusions:** This study demonstrates trends in the published literature on POI and provides new insights for researchers. It emphasizes the importance of multidisciplinary cooperation in the development of this field.

## Introduction

Postoperative ileus (POI), also known as postoperative gastrointestinal dysfunction, frequently occurs after abdominal and other surgical procedures^[1]^. The incidence rate of POI after abdominal surgery ranges from 10 % to 30 %^[2–5]^. The clinical manifestations of POI include feeding intolerance, abdominal distention, abdominal pain, loss of bowel sounds, cessation of flatus and defecation, nausea, and vomiting, all of which impede recovery and impair patients’ quality of life. POI is not only a significant risk factor for complications, such as wound dehiscence, pulmonary embolism, and deep vein thrombosis^[6]^ but also a significant predictor of prolonged hospitalization and increased costs^[7],[8],[9]^. It has been estimated that in the USA, the annual costs associated with POI are as high as US$1.47 billion, placing a significant financial burden on society^[10]^.

Scientometrics reveals the current status and trends of research in a particular field through quantitative analysis of the literature. This helps researchers identify research priorities and development frontiers, which is beneficial for future research^[11],[12]^.

CiteSpace has been used for bibliometric analyses in various research fields; however, bibliometric analyses of POI have not yet been published. In this study, we first used CiteSpace to analyze articles on POI published from 2011 to 2023 based on the Web of Science Core Collection (WoSCC) database to provide guidance for future scientific research on POI.

## Materials and methods

### Search strategies

We extracted literature from the Science Citation Index-EXPANDED and Social Sciences Citation Index databases in the WoSCC via the Chengdu University of Traditional Chinese Medicine Library website. The terms used in the search were TS = (Postoperative ileus OR Postoperative intestinal obstruction OR Postoperative gastrointestinal dysfunction OR Postoperative intestinal paralysis) and Language = English. The time span was limited from January 1, 2011, to May 31, 2023. We selected only original articles and reviews, and all data were downloaded on June 1, 2023. Consequently, 778 records were obtained. S1 Fig shows the details of the search process.

### Analysis tool

In this study, Microsoft Excel 2021, VOSviewer, and CiteSpace 6.2.R2 were selected for the bibliometric analysis, and the data related to countries, institutions, journals, and authors could be generated using the above software. In this study, article count, citation frequency, impact factor (IF), citation/article ratio, centrality, and other parameters were selected. We used article counting to measure productivity and identify productive countries, institutions, journals, and authors. The IF was obtained from the Journal Citation Reports for 2022. IF was used to assess journal quality. We used VOSviewer to build a visual map of the network and examine partnerships between countries and institutions. “Countries” and “organizations” were selected as the units of analysis. The counting method was set to full count. We selected CiteSpace 6.2.R2 to map the keyword co-occurrence view, keyword clustering, and reference citation clustering. The parameters were set as follows: For articles published from 2011 to 2023, the time slice was set to 1 year. The node selection type was set to 1 at a time, with a g-index k value of 5. Different parameters were set according to the different node types to draw visualized maps.

## Results

### Output trends of publications

From 2011 to 2023, 778 POI-related records met the search criteria, including 646 articles and 132 reviews. Fig 1A depicts the annual global distribution and cumulative number of publications on POI from 2011 to 2023. Between 2011 and 2023, the cumulative annual publication trend was linear. Fig 1B illustrates the annual publication trends for the top 10 academic output countries. Over the past decade, the number of publications on POI in China and the USA has significantly increased.

**Fig 1.**
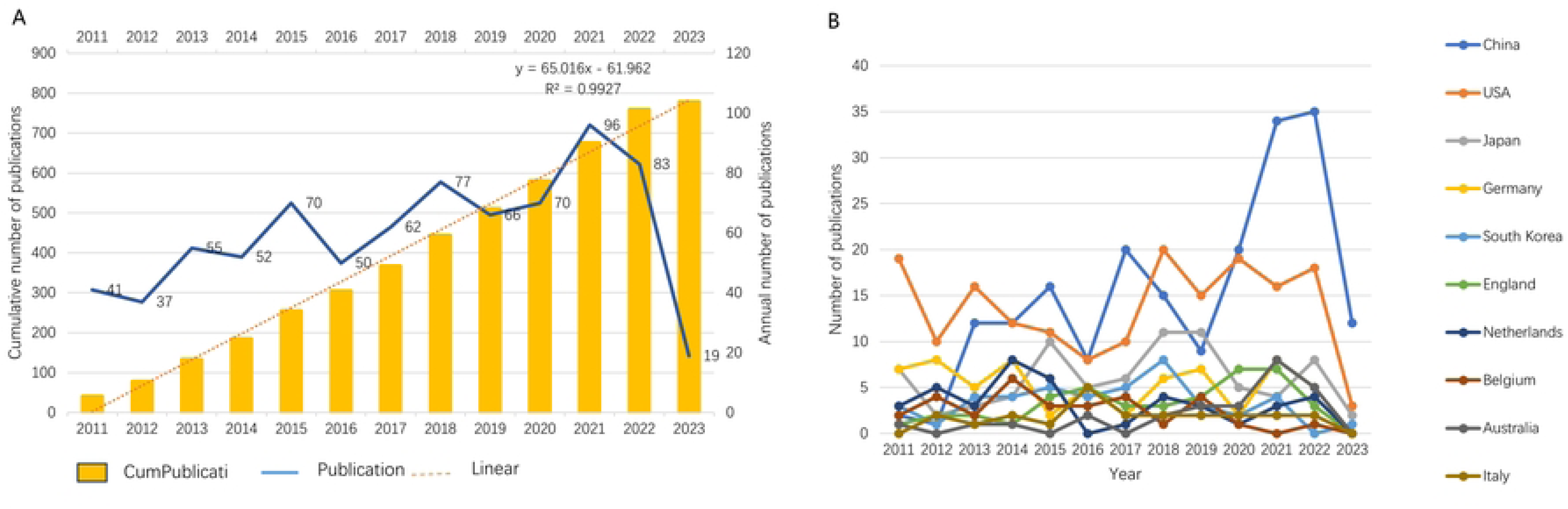
Publication growth trends and analysis of countries in the postoperative ileus field. A: The number of global annual publications and the cumulative number of annual publications; B: Annual publication growth in the top 10 countries.

### Contribution by country and institution

The 778 articles came from 1097 institutions in 59 countries. Table 1 presents detailed information on the top 10 countries. China had the most articles (201), followed by the USA (177) and Japan (78). However, the top three countries in terms of total citations were the USA, China, and the Netherlands. Among the top 10 countries, Belgium showed high academic quality with a significantly higher citation/article ratio (40.55) than the other countries.

**Table 1.**
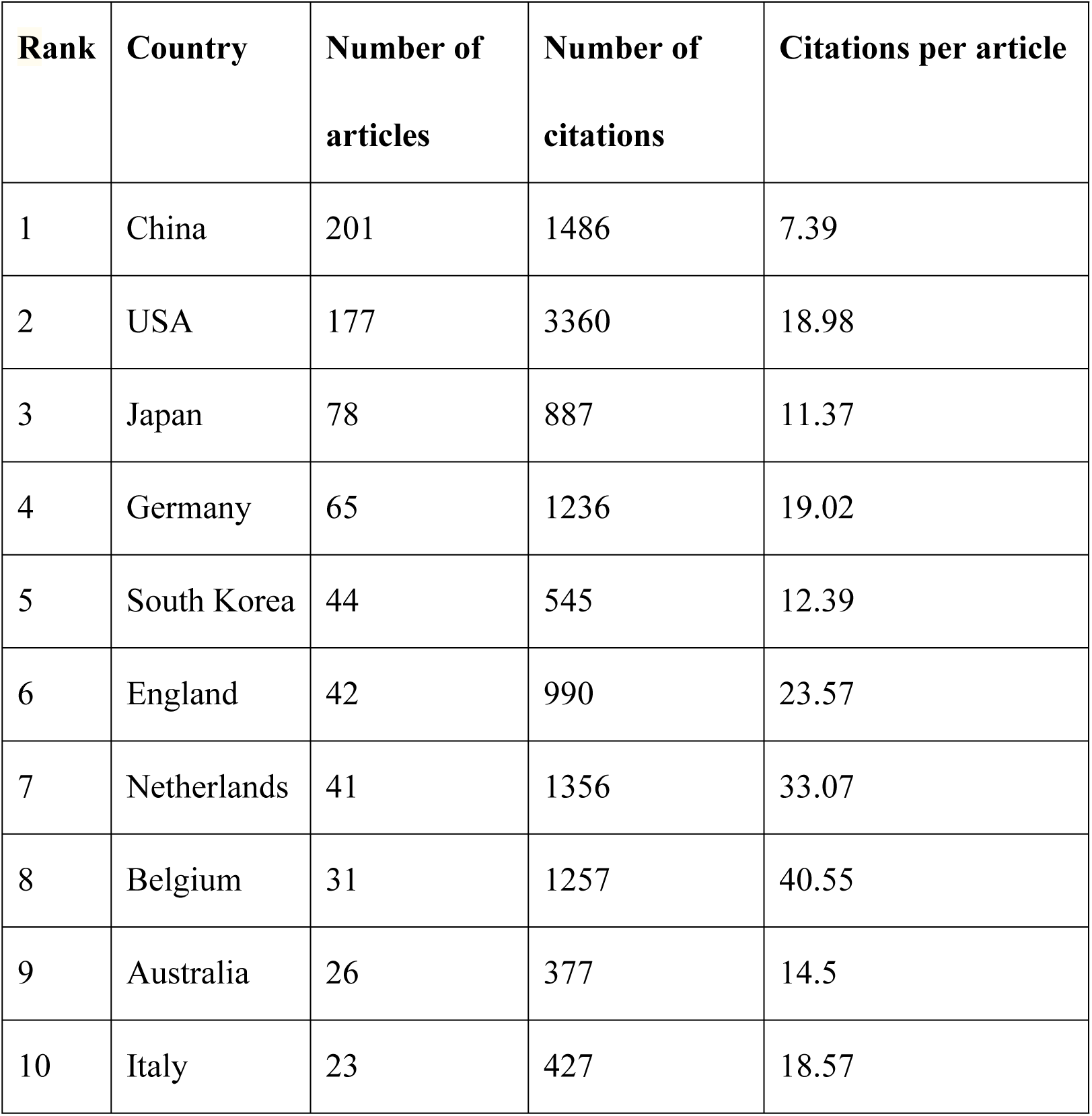
The top 10 countries that published articles on postoperative ileus research.

| **Rank** | **Country** | **Number of**<br>**articles** | **Number of**<br>**citations** | **Citations per article** |
| --- | --- | --- | --- | --- |
| 1 | China | 201 | 1486 | 7.39 |
| 2 | USA | 177 | 3360 | 18.98 |
| 3 | Japan | 78 | 887 | 11.37 |
| 4 | Germany | 65 | 1236 | 19.02 |
| 5 | South Korea | 44 | 545 | 12.39 |
| 6 | England | 42 | 990 | 23.57 |
| 7 | Netherlands | 41 | 1356 | 33.07 |
| 8 | Belgium | 31 | 1257 | 40.55 |
| 9 | Australia | 26 | 377 | 14.5 |
| 10 | Italy | 23 | 427 | 18.57 |

We used VOSviewer to construct network visualization graphs for POI-related articles to assess international cooperation. Collaboration between countries and institutions is shown in S2 Fig. Nodes with a higher degree of co-occurrence were classified as having the same color. Nodes with similar colors formed clusters, indicating closer cooperation. The width of the lines indicated the strength of the cooperation. As shown in the S2 Fig, the USA had the highest total link strength, indicating that it cooperates the most with the rest of the world. The country with which the USA cooperated the most was the Netherlands, followed by Germany and Belgium, and the top 10 most productive institutions are listed in Table 2. The University of Bonn (23 articles) ranked first, followed by the Nanjing University (18 articles), and the University of Tokyo (16 articles). However, the University of Amsterdam had the highest citation/article ratio (55), followed by the University of Auckland (38.14) and Katholieke Universiteit Leuven (29.45). The light blue cluster was led by the Royal Adelaide Hospital, which had the most collaborations with the University of Adelaide. These findings provide useful information regarding outstanding research teams in the field and the establishment of collaborations.

**Table 2.**
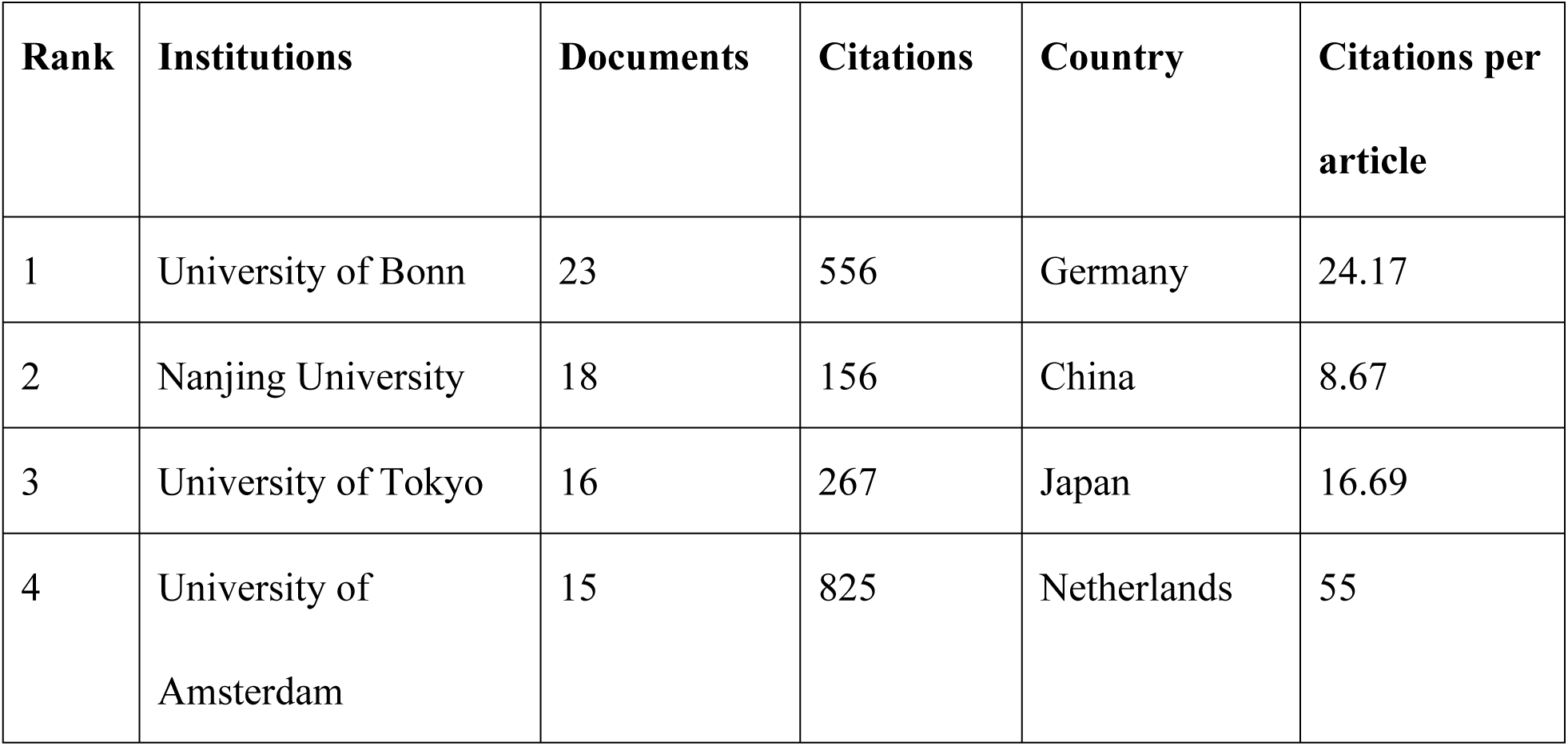
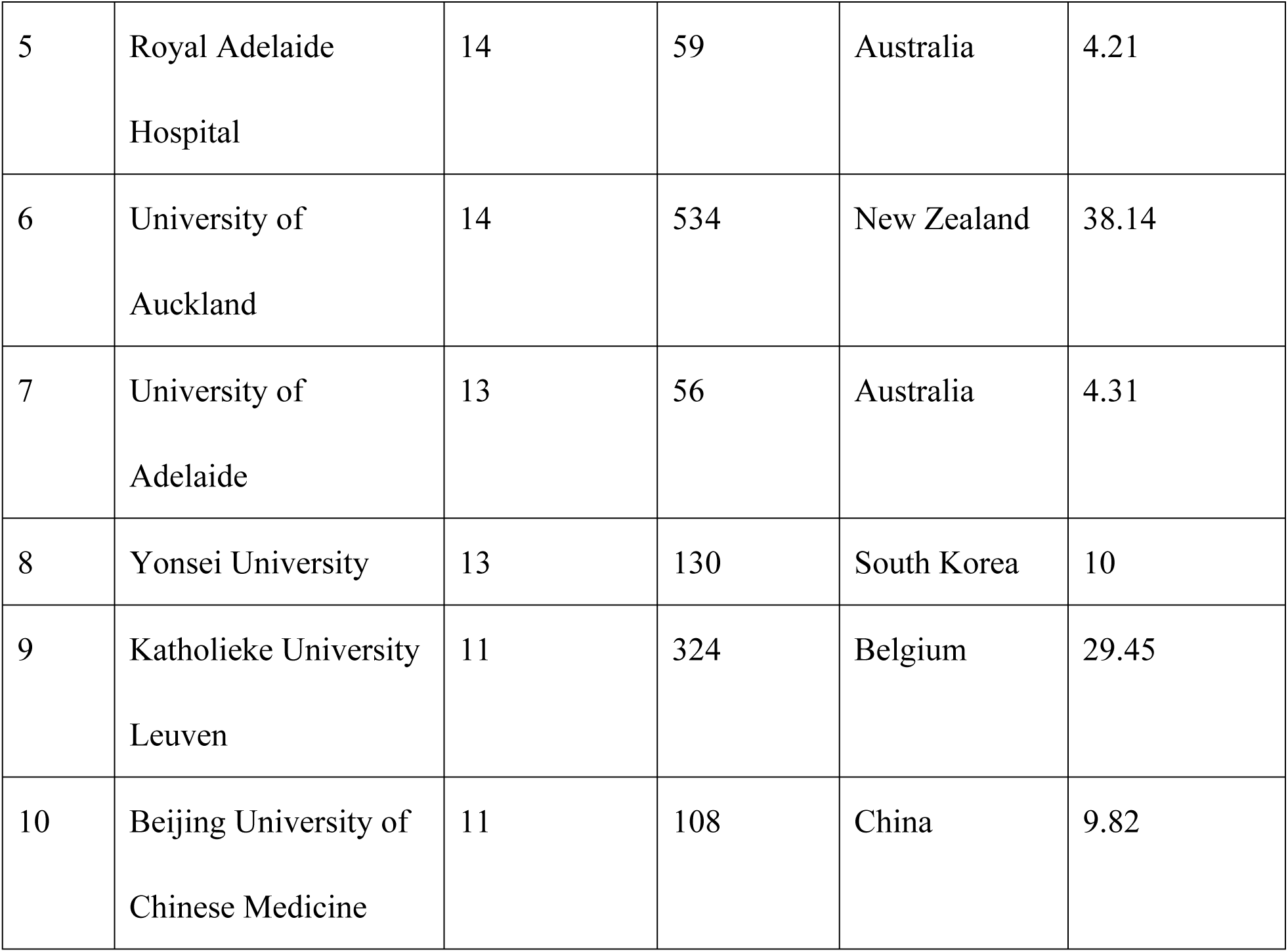
The top 10 institutions that published articles on postoperative ileus.

| Rank | Institutions | Documents | Citations | Country | Citations per article |
| --- | --- | --- | --- | --- | --- |
| 1 | University of Bonn | 23 | 556 | Germany | 24.17 |
| 2 | Nanjing University | 18 | 156 | China | 8.67 |
| 3 | University of Tokyo | 16 | 267 | Japan | 16.69 |
| 4 | University of Amsterdam | 15 | 825 | Netherlands | 55 |
| 5 | Royal Adelaide<br>Hospital | 14 | 59 | Australia | 4.21 |
| 6 | University of<br>Auckland | 14 | 534 | New Zealand | 38.14 |
| 7 | University of<br>Adelaide | 13 | 56 | Australia | 4.31 |
| 8 | Yonsei University | 13 | 130 | South Korea | 10 |
| 9 | Katholieke University<br>Leuven | 11 | 324 | Belgium | 29.45 |
| 10 | Beijing University of<br>Chinese Medicine | 11 | 108 | China | 9.82 |

The labels on the left side of the map represent the disciplines covered by citing journals, and the labels on the right side of the map represent the disciplines covered by cited journals. Colored paths illustrate citation relationships, and thick lines represent major paths.

### Analysis of journals and cited journals

POI research papers were published by 4539 authors in 304 journals, with 15,389 publications in 3384 journals. The top 10 journals accounted for 22.6 % of the total number of articles (778) (S1 Table). Among these journals, *Neurogastroenterology & Motility* published the highest number of articles on POI research (31 articles, IF 2022 = 3.96), followed by the *International Journal of Colorectal Disease* (24 articles, IF 2022 = 2.796). The *British Journal of Surgery* had the highest IF (11.122), and *Journal of Gastrointestinal Surgery* had a citation/article ratio (29.52) that far exceeded that of the other listed journals. S2 Table lists the top 10 journals cited in POI research. *Annals of Surgery* was the most cited journal (1356 citations) in the field, followed by *Diseases of the Colon and Rectum* (798 citations) and *British Journal of Surgery* and *Gastroenterology* (773 citations). Additionally, Fig 2 shows a dual-map overlay of journals reflecting the subject distribution of academic journals. Most clusters of cited journals were located in medicine, medical, clinical, molecular, biology, genetics, health, and nursing.

**Fig 2.**
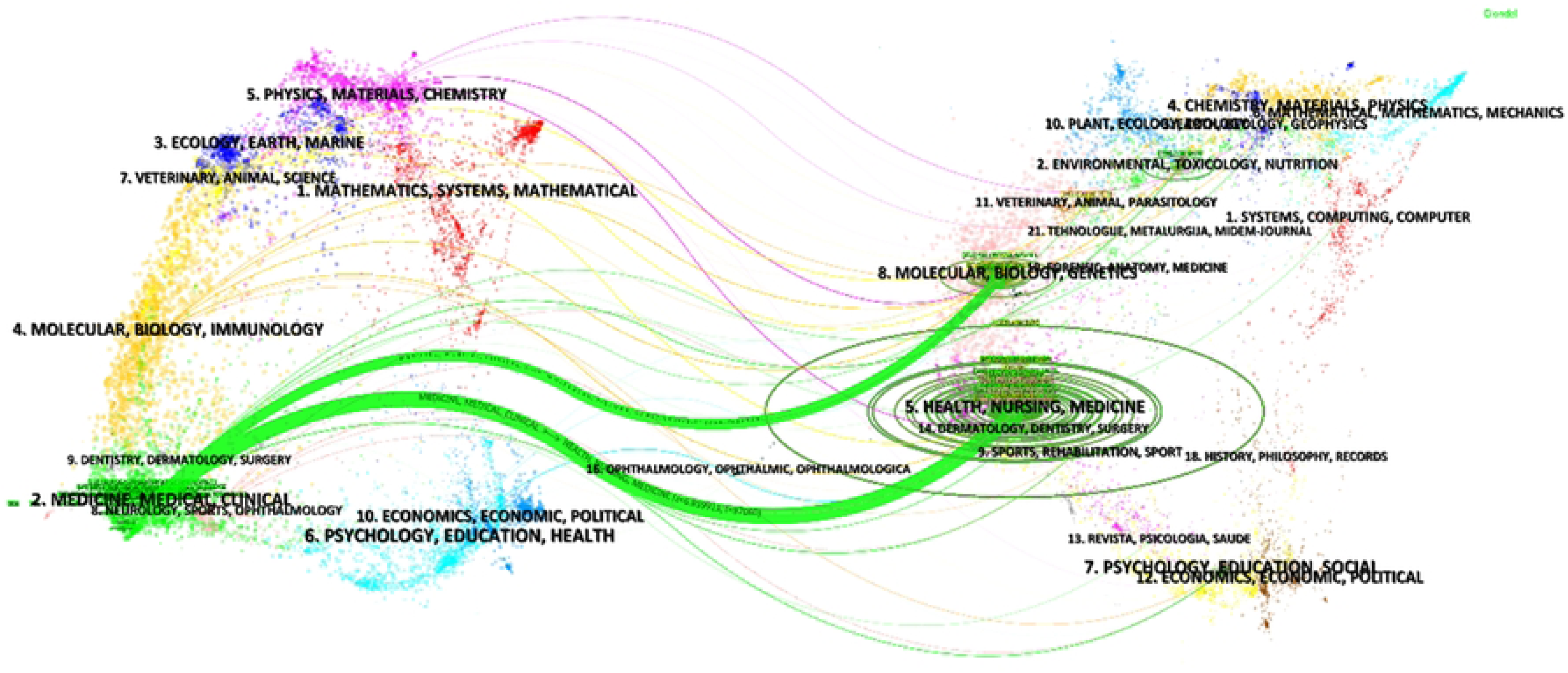
The dual-map overlay of journals in postoperative ileus research.

### Contribution by authors

A total of 4539 authors made contributions to the total number of publications. The most productive authors are listed in S3 Table. Wehner Sven published 22 articles and ranked first in terms of the number of publications, followed by Kalff Joerg (20). Matteoli Gianluca had the highest citation/article ratio (66.78), followed by De Jonge Wouter (32.78) and Wehner Sven (22.27).

### Analysis of keywords

High-frequency and high-centrality keywords reflect the current research hotspots in a particular field^[13]^. We used CiteSpace to create a co-occurrence graph for the 110 keywords (Fig 3A). Table 3 lists the top 10 keywords according to their frequency and centrality. The five most frequent keywords were “postoperative ileus,” “colorectal surgery,” “surgery,” “risk factors,” and “management.” The top 5 keywords with the most centrality were “section,” “enhanced recovery,” “alvimopan,” “double blind,” and “management.” Four of the top 10 most frequent keywords were related to surgery, and two were related to recovery.All the keywords were divided into nine clusters based on a co-occurrence map (Fig 3B). The top five clusters were “inflammation,” “surgery,” “risk factors,” “intestinal obstruction,” and “small bowel obstruction,” showing the structural system of the field of POI research.

**Fig 3.**
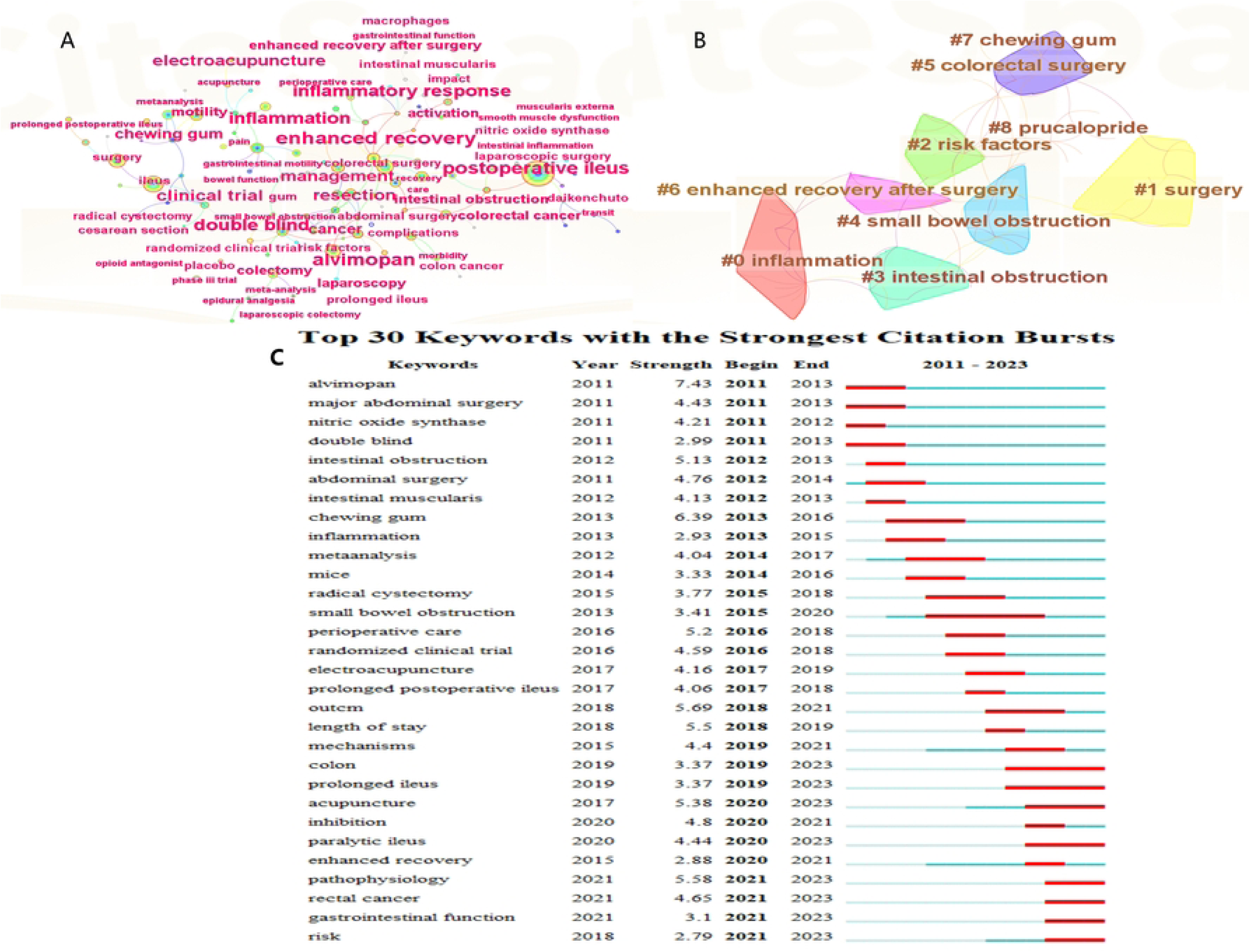
The keywords analysis on postoperative ileus research. A: Co-occurrence map of keywords for postoperative ileus; B: Clusters of keywords in postoperative ileus research; C: Top 30 keywords with the strongest citation bursts on postoperative ileus research.

**Table 3.** Top 10 Keywords with the Highest Frequency and Centrality Values for postoperative ileus.

| Ranking | Frequency | Keyword | Centrality | Keyword |
| --- | --- | --- | --- | --- |
| 1 | 341 | Postoperative ileus | 0.73 | Resection |
| 2 | 120 | Colorectal surgery | 0.65 | Enhanced recovery |
| 3 | 116 | Surgery | 0.62 | Alvimopan |
| 4 | 110 | Risk factors | 0.46 | Double blind |
| 5 | 101 | Management | 0.37 | Management |
| 6 | 80 | Recovery | 0.35 | Postoperative ileus |
| 7 | 77 | Enhanced recovery | 0.35 | Abdominal surgery |
| 8 | 66 | Resection | 0.32 | Intestinal obstruction |
| 9 | 61 | Abdominal surgery | 0.3 | Mechanisms |
| 10 | 58 | Metaanalysis | 0.3 | Inflammatory response |

Burst keywords appear as high-frequency words within a certain time period, showing research hotspots and trends and possibly predicting future emerging focus in dynamic research fields^[14]^. Fig 3C shows the top 30 keywords with the strongest citation bursts from 2011 to 2023. Of these, “alvimopan” had the highest burst strength (7.43) since 2011, followed by “chewing gum” (6.39). “Small bowel obstruction” had the longest burst period (2015–2020). The latest burst keywords were “colon” (2019– 2023), “prolonged ileus” (2019–2023), “acupuncture” (2020–2023), “paralytic ileus” (2020–2023), “pathophysiology” (2021–2023), “rectal cancer” (2021–2023), “gastrointestinal function” (2021–2023), and “risk” (2021–2023), indicating that they have been the focus of attention so far.

### Analysis of references

Co-cited references highlight key articles that contribute to research in the field. S4 Table lists the top 10 highly cited references over the past 12 years. The milestone literature and rapid evolution of the field can be elucidated by citation analysis of relevant documents^[15]^. Productions with the strongest citation bursts are considered the knowledge bases of the research frontiers. We utilized CiteSpace to generate a timeline visualization of the cited references (Fig 4A), clearly showing the development trends of POI research. The largest cluster was “coffee”, indicating a significant research focus and interest over the past several years. This was followed by “prolonged postoperative ileus”, “chewing gum”, “mast cells”, “postoperative care”, and “gut microbiota”. Clusters 2 “chewing gum”, 3 “mast cells”, 4 “postoperative care”, 10 “alvimopan”, and 11 “ghrelin” received more attention in the early years, and clusters 0, 1, 5, 9, and 12 continue to evolve, foreshadowing future research trends.

**Fig 4.**
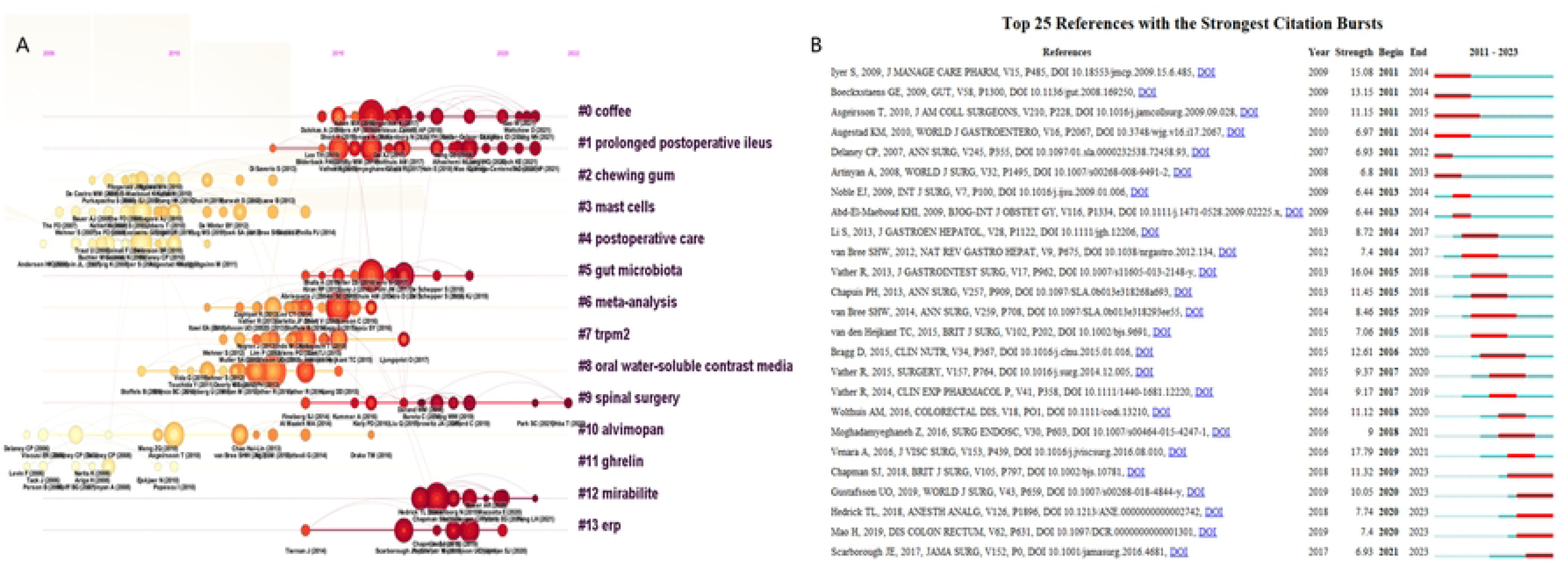
The references analysis on postoperative ileus research. A: Timeline view of the cited references in postoperative ileus research. B: Top 25 references with the strongest citation bursts on postoperative ileus research

Fig 4B shows the top 25 references with the strongest citation bursts. Citation bursts until 2023 were led by Chapman et al. (2018), who had the strongest burst (11.32), followed by Gustafsson et al. (2019), Hedrick et al. (2018), Mao et al. (2019), and Scarborough et al. (2017). References on the definition, pathophysiologic mechanisms, prevention, and financial impact of POI demonstrated active citation bursts^[16–19]^.

## Discussion

In this study, we performed a bibliometric analysis of documents related to POI from 2011 to 2023 by searching the WOSCC. The aim was to summarize the current state of research, hotspots, and trends in this field from a global perspective.

In total, 778 documents on POI were retrieved, including 646 articles and 132 reviews. From 2011 to 2023, the number of annual articles in this field was generally on the rise, indicating that research on POI is increasing annually. China was the most productive country, especially between 2019 and 2021, with rapid growth in the number of publications. Nevertheless, although China had the highest number of articles, its citation/article ratio ranked last among the top 10 countries. Therefore, the overall quality of the articles requires further improvement. Therefore, China must focus on in-depth research to further strengthen its cooperation with other countries. In contrast, although Belgium had published only a few articles, these articles had high academic quality.

Of the top 10 institutions, nine were universities, indicating that universities are the main research community in this field. Two of the top 10 institutions were from China (Nanjing University and Beijing University of Chinese Medicine), and two were from Australia (University of Adelaide and Royal Adelaide Hospital), suggesting that these two countries have a multitude of excellent research groups in this field. The IF scores of the retrieved articles of the top 10 most productive journals were generally low. Therefore, high-quality research in this field is required.

Furthermore, the double map overlay of the journals shows that this study concentrated on clinical medicine and basic research; consequently, multidisciplinary cross-collaboration is required in the future to promote in-depth development of the POI domain.

This study has identified several excellent research outputs that have contributed significantly to the advancement of POI research. For example, (Vather R, 2013)^[20]^was most widely cited. It proposed clinically quantifiable definitions of POI and standardized clinical parameters that can help to objectively compare different intervention modalities and were valuable for the improvement of external validity of future clinical trials. Chapuis et al.^[4]^ identified independent preoperative and intraoperative risk factors for prolonged ileus, which will not only help healthcare professionals to identify suspected patients and provide targeted treatment, but will also help researchers more effectively select subjects of clinical trials for the prevention or treatment of prolonged ileus. Ng et al.^[21]^ conducted a randomized controlled trial on electroacupuncture for POI and confirmed that electroacupuncture reduced the duration of ileus, time to first out-of-bed activity, and need for postoperative analgesia after laparoscopic surgery for colorectal cancer. This study provides high-quality evidence for the efficacy of acupuncture for the treatment of POI.

Analysis of keyword bursts can assist in predicting research hotspots and frontiers in POI areas. The keywords “colon,” “prolonged ileus,” “acupuncture,” “paralytic ileus,” “pathophysiology,” “rectal cancer,” and “gastrointestinal function” are anticipated to appear frequently over the next few years, indicating emerging research trends. The top three POI frontiers are as follows: (1) prolonged ileus: prolonged ileus is usually called paralytic ileus, which is defined as the absence of bowel function recovery within 7 days after surgery^[5]^. Prolonged ileus is a common complication of colon surgery, with an incidence rate of up to 12.7%, and is significantly associated with higher mortality, longer postoperative hospital stay, and higher risks of unplanned reoperation and readmission^[5]^. The independent risk factors for prolonged ileus are male sex, respiratory diseases, peripheral vascular disease, resection at urgent operation, transfusion in perioperative period, construction of stoma, and operation lasting ≥ 3 h^[4]^. Such patients are more prone to postoperative complications, such as intra-abdominal infection and anastomotic leakage^[2, 5]^. Prolonged ileus treatment is estimated to cost 750 million dollars per year in the USA ^[22]^. A systematic review suggested the selective use of nasogastric tubes to manage prolonged ileus^[23]^; however, new strategies for the treatment of prolonged ileus require further research, as reliable evidence is still lacking^[1]^. (2) Acupuncture: A previous study reported the increasing popularity of acupuncture in the field of POI^[24]^. Animal studies^[25, 26]^ have confirmed that electroacupuncture at ST-36 can accelerate colonic motility and promote colonic contraction through the parasympathetic nervous system and cholinergic pathway. Several clinical studies^[21, 27, 28]^ have found that electroacupuncture reduces the duration of POI after laparoscopic colorectal cancer surgery, time to get out of bed, and need for postoperative analgesics and reduces the incidence of prolonged ileus. The somatic autonomous reflex allows electroacupuncture to regulate physiological functions at specific acupoints^[29]^. Evidence suggests that electroacupuncture regulates gastrointestinal function by stimulating the vagus nervous system and may produce anti-inflammatory effects^[30, 31]^. An experimental study revealed that electroacupuncture activated the vagus nerve, which in turn activated the nicotinic alpha 7 acetylcholine receptors (α7nAChR)-mediated JAK2/STAT3 signaling pathway in macrophages, thereby inhibiting intestinal operation-induced inflammation^[32]^. However, some researchers have found that the regulatory effect of electroacupuncture on POI is mainly achieved by stimulating nucleus tractus solitarii neurons to promote gastrointestinal motility but not by activating the cholinergic anti-inflammatory pathway^[33]^. Therefore, the exact mechanism of action of acupuncture for the treatment of POI requires further investigation. (3) Pathophysiology: POI is caused by autonomic and hormonal mechanisms^[19]^. The triggers of POI are multifactorial because bowel manipulation, fluid overload, neurohormonal dysfunction, inflammation, opioid analgesia, and somatic and visceral trauma are all associated with the condition^[1, 34–37]^. POI consists of three stages of development. The first stage of skin incision involves the sympathetic nervous system and is mediated by the corticotropin-releasing factor, leading to acute intestinal paralysis^[37]^. The second stage begins 3–4 h after intestinal manipulation and is mediated by inflammation. The release of local inflammatory cytokines and chemokines upregulates the expression of adhesion molecules in endothelial cells^[7]^. Consequently, leukocytes, mainly monocytes and neutrophils invade the muscularis externa of the small intestine^[38]^. Invading mononuclear cells and activated macrophages release nitric oxide and prostaglandins, which inhibit smooth muscle contraction^[39]^. Local inflammation leads to widespread suppression of gastrointestinal motodynamics through a “field effect,”^[40]^ whose mechanisms include complex neuronal and immune responses, including white blood cell production of nitric oxide^[39]^, whole-intestinal dissemination of T helper cell-mediated inflammation^[39]^, and activation of inhibitory neural pathways affecting the entire intestine^[41]^. The invasion of leukocytes into the muscularis externa (ME) underlies the impairment of intestinal motility after intestinal manipulation^[38]^. However, the triggers of the inflammatory cascade are unknown and may involve dendritic cells, mast cells, and/or macrophages^[6]^. The third phase involves the activation of the vagus nerve system. It is mediated by the α7nAChR and 5-hydroxytryptamine 4 receptors (5-HT4R)^[19]^. Activation of 5-HT4R leads to an increase in acetylcholine released by myenteric cholinergic neurons, thereby activating α7nAChR on monocytes and macrophages and reducing the inflammatory response^[42]^. The development of the POI includes the enteric gliosis of the ME^[43]^. The latest cutting-edge research^[44]^ indicated that beta adrenergic signaling is a priming factor in triggering enteric glial responsiveness and the onset of acute enteric gliosis during surgical procedures, which presents that beta adrenergic signaling of enteric glia is a promising target for averting the emergence of POI. Most of the aforementioned studies on the pathophysiological mechanisms of POI have involved mouse models. However, data obtained from mouse models may not be applicable to humans. For example, the expression profiles of mediators and receptors differ between humans and rodents^[45]^. It is difficult to generate animal models of complications such as diabetes and hypertension^[6]^. Therefore, further clinical studies are required to elucidate the mechanisms underlying POI in humans.

Articles with the highest burst of citations are potentially valuable for exploring research frontiers^[11]^. In this study, “coffee” was the largest cluster, and ^“^spinal surgery^”^ was active until the most recent publication year. There is no consensus on whether coffee can promote the recovery of POI. Randomized controlled trial (RCT) and meta-analysis showed that coffee was effective in shortening postoperative defecation time and hospital stay, which can be used to prevent POI ^[46–48]^. However, a RCT^[49]^ suggested that coffee could not accelerate the recovery of intestinal function after minimally invasive surgery. Therefore, the study of coffee in the treatment of POI may be a research hotspot in the future. POI is a common complication of spinal surgery, with an incidence rate of 5–12%^[50]^. It is expected that there will be a large number of future studies on POI after spinal surgery. The top three publications with the strongest citation bursts were as follows: (1) Chapman et al.^[16]^ systematically reviewed the pathophysiological mechanisms of POI. Current pathophysiological knowledge acknowledges neurogenic, inflammatory, and pharmacological mechanisms. However, most evidence comes from animal studies, and the exact physiological mechanism of POI in humans requires further study. Furthermore, they found that the methodological preconditions for POI research are lacking. This study provides a reference for future POI researches. (2) Gustafsson et al.^[51]^coauthored guidelines that provide consensus on optimal perioperative care for colorectal surgery and provide evidence-based grading recommendations for each enhanced recovery after surgery (ERAS) program in the ERAS protocol. These guidelines strongly recommend multimodal prevention of POI. (3) Hedrick et al.^[18]^ published a joint consensus statement for the second Perioperative Quality Initiative, which reached a consensus on the definition of POI and established a framework for future clinical and scientific work. To provide consensus recommendations on current prevention strategies in view of the extensive postoperative impairment of gastrointestinal function, reasonable countermeasures were formulated.

## Conclusions

This study presents the global trends, keywords, highly cited articles, and research frontiers in the POI field from 2011 to 2023. According to the obtained data, POI research is in the rising stage of development and has great growth potential. China leads the way in the number of publications. Research hotspots in the field of POI have mainly focused on prolonged ileus, acupuncture, and its pathophysiology. In the future, it will be necessary to further strengthen the cooperation between countries and institutions, especially to pay attention to the importance of multidisciplinary cooperation in the development of this field.

## Data Availability

All relevant data are within the manuscript and its Supporting Information files.

## Acknowledgments

We would like to thank Editage (www.editage.com) for English language editing.

## Supporting information

**S1 Fig. A flowchart representing retrieval strategies for postoperative ileus articles from the Web of Science Core Collection database.**

**S2 Fig. Scientific influence in the field of postoperative ileus worldwide. A: Network plot of influential countries among the publications of Web of Science Core Collection (WoSCC); B: Network plot of influential institutions among the publications of WoSCC.**

**S1 Table. The top 10 journals that published articles on postoperative ileus research.**

**S2 Table. The top 10 cited journals of postoperative ileus research.**

**S3 Table. The top 10 authors that published articles on postoperative ileus research.**

**S4 Table. Top 10 highly cited publications in postoperative ileus.**

